# Implantable loop recorder in diagnostics of cardiac arrhythmias: the importance of drug treatment in predicting pacemaker requirement

**DOI:** 10.1101/2025.04.18.25326090

**Authors:** Jelena Vuckovic-Filipovic, Violeta Iric Cupic, Goran Davidovic, Vladimir Ignjatovic, Vesna Ignjatovic, Isidora Stankovic, Ivan Simic, Vladimir Miloradovic, Natasa Djordjevic

**Author notes:** **Corresponding:** Vladimir Ignjatovic, MD, PhD, Department of Internal Medicine, Faculty of Medical Sciences, University of Kragujevac, Svetozara Markovica 69, 34000 Kragujevac, Serbia.

## Abstract

**Background:** Implantable loop recorder (ILR) represents gold standard in diagnosis of cardiac arrhythmias in patients presenting with transitory neurological or cardiac symptoms. The aim of our study was to determine its real-world diagnostic effectiveness detecting arrhythmias requiring pacemaker implantation.

**Methods:** This retrospective, observational clinical study enrolled 62 ILR recipients from Cardiology Clinic of the Clinical Center Kragujevac, which were followed-up for 2 years. Demographic and clinical data were obtained from patients’ medical records.

**Results:** By the end of the monitoring, some form of cardiac rhythm disorder (most commonly bradyarrhythmias) was observed in 36% of the study population, and in 34% a pacemaker or cardioverter defibrillator was implanted. The most common indication for pacemaker implantation were pauses in cardiac activity (83%), followed by bradycardia (33%); atrioventricular block (11%) and atrial fibrillation (6%). Univariable logistic regression analysis showed that the use of oral anticoagulants (OR [95% CI]: 0.014 [1.77; 154.55]), ACE inhibitors or AT receptor blockers (OR [95% CI]: 0.022 [1.210; 12.352]), and diuretics (OR [95% CI]: 0.008 [1.549; 18.035]) had a statistically significant impact on the diagnostic value of ILR in detecting arrhythmias requiring pacemaker. After adjustment for other factors of influence, only oral anticoagulants (OR [95% CI]: 0.042 [1.091; 1112.8]) and diuretics (OR [95% CI]: 0.043 [1.047; 14.623]) remained significant in predicting pacemaker requirement in ILR recipients.

**Conclusion:** Our study shows that ILR represents an effective diagnostic approach in detecting cardiac arrhythmias requiring pacemaker implantation, especially in patients treated with oral anticoagulants or diuretics. The relevance of previous treatment with ACE inhibitors or AT receptor blockers remains to be confirmed in the future

## Introduction

Cardiac arrhythmias consist of a diverse group of disorders that vary by origin, prevalence, characteristics, and severity, but align in posing a risk of serious complications, such as unexplained syncope (1). To establish symptom-rhythm correlation, several diagnostic procedures have been proposed, including elective short-term electrocardiogram (ECG) and Holter monitoring, as well as provocative head-up tilt and electrophysiological testing (2, 3). Being often asymptomatic and unpredictable in nature, cardiac arrhythmias proved to be difficult to diagnose, making elective tests less useful, and provocative methods less specific (4, 5). However, even rare arrhythmias can be detected by prolonged monitoring, and coincidence between an abnormal ECG finding and a syncopal episode or palpitations can be considered gold standard of diagnosis (4).

Implantable loop recorder (ILR) is a small electronic device that can be inserted subcutaneously in the chest to continuously monitor the cardiac rhythm; it will automatically record and store (in the form of ECG) any significant arrhythmia that occurs during the lifetime of its battery of up to 5 years (6–8). Based on the newest European Stroke Organization guideline, the use of ILR is recommended for detection of subclinical atrial fibrillation as a possible (and treatable) underlying cause of ischemic stroke or transient ischemic attack of undetermined origin (9). Similarly, the European Society of Cardiology (ESC) (10) currently recommends using ILR in diagnostics of arrhythmias in patients with unexplained syncope, especially in the presence of chronic coronary artery disease and reduced ejection fraction, or cardiomyopathy (class I recommendation). Another established indication for ILR are unexplained palpitations (11, 12), where this method represents more efficient and more cost-effective diagnostic approach as compared to other conventional strategies (13, 14). Other less common conditions reported to benefit from ILR application include congenital heart disease, cardiac light chain amyloidosis, obstructive sleep apnea, epilepsy, unexplained falls, and monitoring after coronary artery bypass grafting (15, 16).

The efficacy of ILR in recording concealed cardiac arrhythmias has been demonstrated by several studies. These include recent systematic review and meta-analysis of randomized controlled trials involving 7237 patients (17), which concluded that ILR-based screening leads to increased detection of incident arrhythmias and increased initiation of appropriate drug therapy. However, the real-world data regarding the diagnostic yield of ILR use in different clinical settings seem to be scarce (8), especially in relation to the conditions requiring treatment with interventional cardiology procedures (18). Therefore, the aim of our study was to determine diagnostic effectiveness of ILR in detecting arrhythmias as an indication for pacemaker or cardioverter defibrillator implantation in patients presenting with transitory neurological or cardiac symptoms.

## Methods

This clinical, retrospective, observational study involved patients that have received an implantable loop recorder (ILR) at the Cardiology Clinic of the Clinical Center Kragujevac from January 2019 to January 2022. To be included in the study, patients had to fulfill all the inclusion criteria, as follows: 1) the presence of neurological or cardiac symptoms as an indication for ILR implantation; 2) completed two-year follow-up period, or occurrence of a clinical event that required discontinuation of follow-up and further treatment; 3) availability of complete medical documentation. Subjects younger than 18 years of age, as well as pregnant or breastfeeding women, were not considered eligible. All study participants signed an informed consent form prior to entering the study. The study was approved by the relevant Ethics Committee of the Clinical Center Kragujevac, decision No. 01/25-53.

Demographic and clinical characteristics of the subjects, including gender, age, indication for ILR implantation and associated symptoms, associated cardiovascular and other diseases, chronic therapy, and the result of endocranial computed tomography (CT), were collected from the patients’ medical records. The main study outcome was the presence of arrhythmias detected by ILR as an indication for implantation of a pacemaker or implantable cardioverter-defibrillator (ICD). The statistical program SPSS version 23 was used for statistical data processing. The normality of the distribution of continuous variables was checked by the Kolmogorov-Smirnov test; in case of a normal distribution, the mean and standard deviation (SD) were used as measures of central tendency and variability; otherwise the median and interquartile range (IQR) were employed. Categorical variables were presented as absolute numbers and percentages. The influence of independent variables on the outcomes was analyzed by univariable and multivariable binary logistic regression. The results are presented in text, tabular or graphical form, with a marginal probability value of less than 0.05.

## Results

A total of 62 patients participated in the study, of whom 24 (38.7%) were women and 38 (61.3%) were men. The age structure did not follow a normal distribution (D(62)=0.138; p=0.005), with the median (IQR) of 49.5 (35.8–67.8) years. Indications for ILR implantation included syncope (66%), palpitations (27%), stroke (6%), or fainting (48%), with 27 (44%) patients presenting at least two of them concurrently. Among those in whom ILR was implanted for syncope, prodromal symptoms were present in 19 patients, i.e. 46%.

During the two-year follow-up, syncope occurred occasionally in 43 (69%) patients, while in 21 (49%) these attacks were characterized as frequent. CT of the endocranium was performed in 28 patients, most of whom (93%) had normal findings. Among the respondents, 41 patients (66%) had at least one associated disease (most frequently (61%) arterial hypertension), while 12 (19%) presented with two or more simultaneously. Among participants, 46 (74%) were on chronic therapy with at least one drug, while 34 (55%) used at least two. The prevalence of drug groups with which the subjects were treated is shown by Figure 1, while the types of drugs according to individual groups are enlisted in Table 1.

**Table 1:**
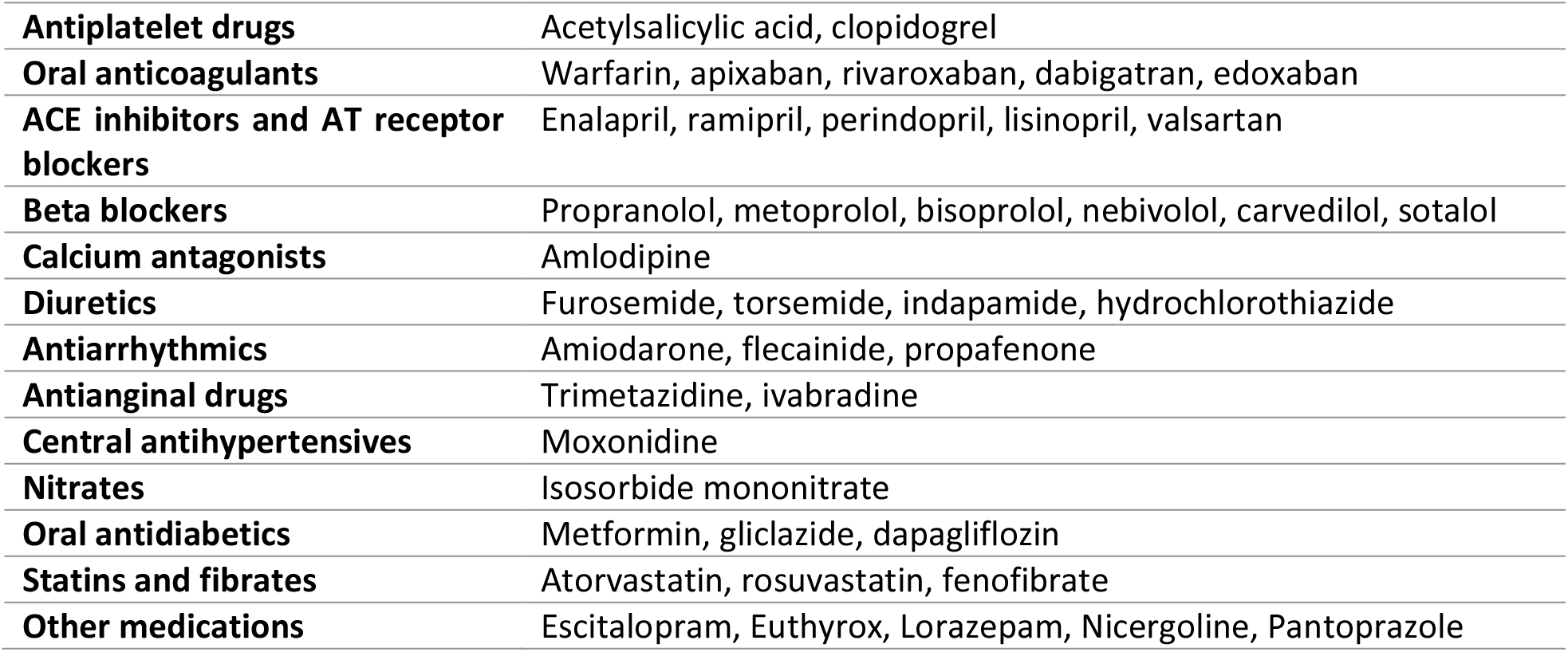
Types of medications used during follow-up.

|  |  |
| --- | --- |
| **Antiplatelet drugs** | Acetylsalicylic acid, clopidogrel |
| **Oral anticoagulants** | Warfarin, apixaban, rivaroxaban, dabigatran, edoxaban |
| **ACE inhibitors and AT receptor blockers** | Enalapril, ramipril, perindopril, lisinopril, valsartan |
| **Beta blockers** | Propranolol, metoprolol, bisoprolol, nebivolol, carvedilol, sotalol |
| **Calcium antagonists** | Amlodipine |
| **Diuretics** | Furosemide, torsemide, indapamide, hydrochlorothiazide |
| **Antiarrhythmics** | Amiodarone, flecainide, propafenone |
| **Antianginal drugs** | Trimetazidine, ivabradine |
| **Central antihypertensives** | Moxonidine |
| **Nitrates** | Isosorbide mononitrate |
| **Oral antidiabetics** | Metformin, gliclazide, dapagliflozin |
| **Statins and fibrates** | Atorvastatin, rosuvastatin, fenofibrate |
| **Other medications** | Escitalopram, Euthyrox, Lorazepam, Nicergoline, Pantoprazole |

**Figure 1:**
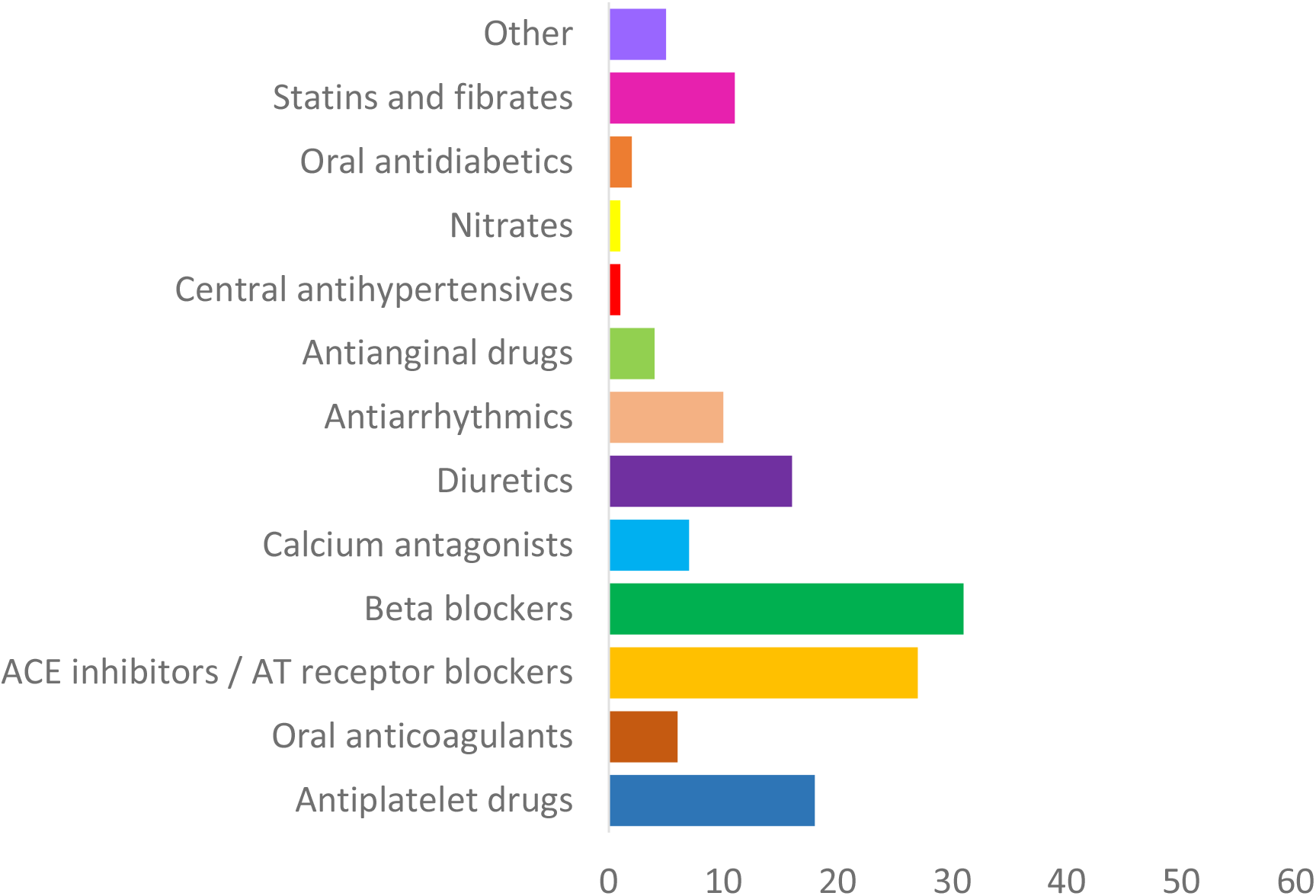
The prevalence of drug groups used by the study population.

By the end of the monitoring, coronary artery disease was diagnosed in 10 subjects (16%), hypertrophic myocardiopathy in 6 (10%), and isolated aortic stenosis in 2 (3%) patients. Based on the electrocardiographic recording by ILR, in 24 patients, which made up 36% of the study population, some form of cardiac rhythm disorder was observed, of which bradyarrhythmias were the most common (Figure 2). Bradycardia, pauses in cardiac activity or atrioventricular block were observed in a total of 20 subjects (32%), of whom 11 (18%) had at least two different arrhythmias detected. Tachyarrhythmias such as ventricular tachycardia, ventricular extrasystoles or atrial fibrillation were significantly less common: at least one of them was observed in only 4 patients (6%), while more than two were detected in only one of them.

**Figure 2:**
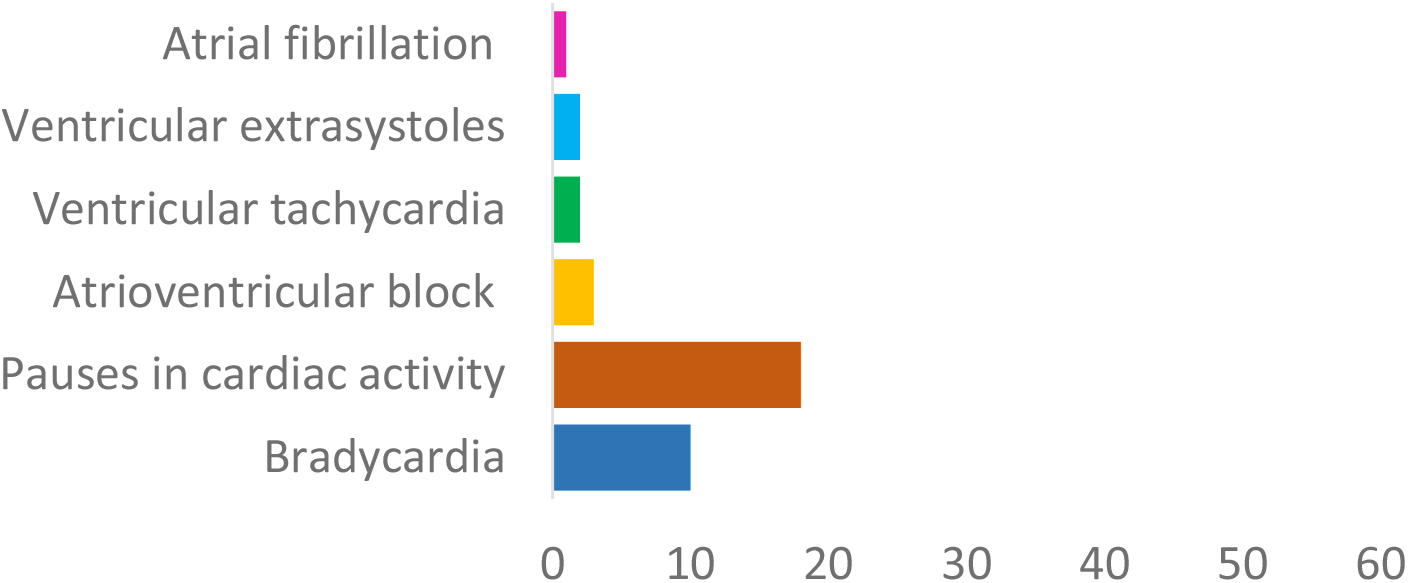
The prevalence of cardiac rhythm disorders in the study population.

Due to arrhythmia detected by ILR, in a total of 21 (34%) patients a pacemaker or cardioverter defibrillator was implanted. The distribution of cardiac rhythm disorders as an indication for the above procedures is shown in Figure 3. The most common indication for pacemaker implantation were pauses in cardiac activity (83%), followed by bradycardia (33%); atrioventricular block (11%) and atrial fibrillation (6%) were significantly less common (some of the patients presented with more than one type of arrhythmia). An implantable cardioverter defibrillator was inserted in only 3 patients.

**Figure 3:**
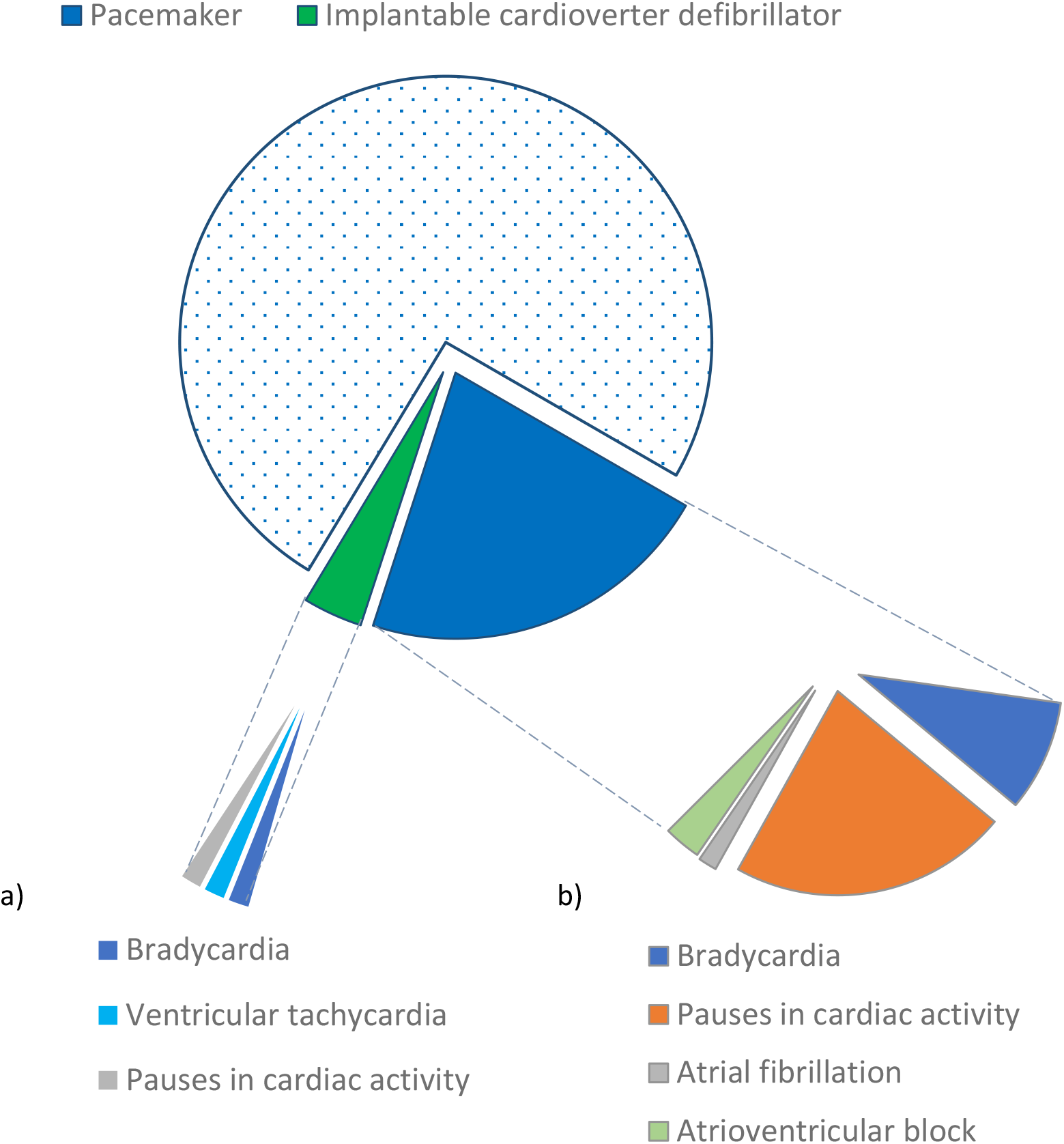
Cardiac rhythm disorders as indications for implantation of a) pacemaker or b) an implantable cardioverter defibrillator in the study population.

Univariable logistic regression analysis showed that only chronic use of oral anticoagulants, ACE inhibitors or AT receptor blockers, and diuretics had a statistically significant impact on the diagnostic value of ILR in determining the presence of an indication for pacemaker implantation (Table 2). Statistical analysis of the influence of factors on the diagnostic value of ILR in determining indications for implantable cardioverter defibrillator implantation was not conducted due to the small sample size.

**Table 2.** Parameters of univariable analysis of the influence of factors on the diagnostic value of ILR in determining indications for pacemaker implantation.

| | B | SE | Wald $\chi^2$ | df | p | OR | 95% CI |
| --- | --- | --- | --- | --- | --- | --- | --- |
| **Demographic characteristics** |  |  |  |  |  |  |  |
| Gender | 0.325 | 0.587 | 0.308 | 1 | 0.579 | 1.385 | 0.439-4.371 |
| Age | 0.033 | 0.018 | 3.489 | 1 | 0.062 | 1.033 | 0.998-1.070 |
| **Indications for ILR** |  |  |  |  |  |  |  |
| Syncope | 0.396 | 0.612 | 0.418 | 1 | 0.518 | 1.486 | 0.447-4.935 |
| Prodromes | 0.539 | 0.574 | 0.881 | 1 | 0.348 | 1.714 | 0.556-5.283 |
| Palpitations | -0.384 | 0.656 | 0.342 | 1 | 0.559 | 0.681 | 0.188-2.466 |
| Stroke | -0.218 | 1.190 | 0.034 | 1 | 0.855 | 0.804 | 0.078-8.285 |
| Dizziness | 0.405 | 0.563 | 0.519 | 1 | 0.471 | 1.500 | 0.498-4.519 |
| **Associated diseases** |  |  |  |  |  |  |  |
| Arterial hypertension | 0.681 | 0.608 | 1.255 | 1 | 0.263 | 1.976 | 0.600-6.505 |
| Diabetes | -0.721 | 0.838 | 0.741 | 1 | 0.389 | 0.486 | 0.094-2.512 |
| Anemia | 0.928 | 1.443 | 0.414 | 1 | 0.520 | 2.529 | 0.150-42.791 |
| Epilepsy | 22.154 | 40,192.9 | 0.000 | 1 | 1.000 | / | / |
| Other hereditary diseases | -20.38 | 23,205.4 | 0.000 | 1 | 0.999 | / | / |
| **Chronic therapy** |  |  |  |  |  |  |  |
| Antiplatelet drugs | -0.950 | 0.708 | 1.802 | 1 | 0.180 | 0.387 | 0.097-1.549 |
| Oral anticoagulants | 2.806 | 1.140 | 6.054 | 1 | **0.014** | 16.538 | 1.770-154.550 |
| ACE inhibitors and AT receptor blockers | 1.352 | 0.593 | 5.209 | 1 | **0.022** | 3.867 | 1.210-12.352 |
| Beta blockers | -0.314 | 0.562 | 0.312 | 1 | 0.576 | 0.730 | 0.243-2.199 |
| Calcium antagonists | -0.025 | 0.888 | 0.001 | 1 | 0.977 | 0.975 | 0.171-5.555 |
| Diuretics | 1.665 | 0.626 | 7.070 | 1 | **0.008** | 5.286 | 1.549-18.035 |
| Antiarrhythmics | 0.593 | 0.717 | 0.684 | 1 | 0.408 | 1.810 | 0.444-7.380 |
| Antianginal drugs | -20,404 | 20,096.5 | 0.000 | 1 | 0.999 | / | / |
| Central antihypertensives | -20.332 | 40,192.9 | 0.000 | 1 | 1.000 | / | / |
| Nitrates | -20.332 | 40,192.9 | 0.000 | 1 | 1.000 | / | / |
| Oral antidiabetics | -20.356 | 28,420.7 | 0.000 | 1 | 0.999 | / | / |
| Statins and fibrates | 0.412 | 0.701 | 0.346 | 1 | 0.556 | 1.510 | 0.382-5.966 |
| Other medications | 1.435 | 0.961 | 2.229 | 1 | 0.135 | 4.200 | 0.638-27.630 |

Multivariable logistic regression with stepwise elimination showed that, when the model was adjusted for the influence of other variables, only the use of oral coagulants and diuretics retained statistical significance (Table 3).

**Table 3.**
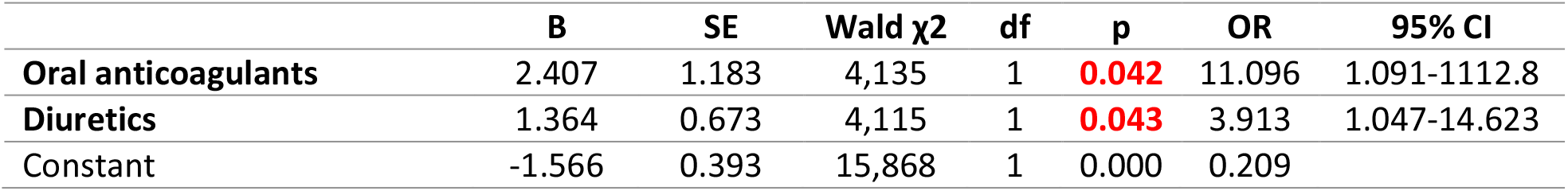
Parameters of multivariable analysis of the influence of factors on the diagnostic value of ILR in determining indications for pacemaker implantation.

The final model showed statistical significance (χ2(2)=12.647; p=0.002), explained 26.4% of the variance in the outcome, and correctly classified 77.4% of cases. The result indicates that patients on oral anticoagulants or diuretics are more likely to be diagnosed by ILR with arrhythmias as an indication for pacemaker implantation.

## Discussion

In the present study, more than one third of patients with inserted ILR due to transitory neurological or cardiac symptoms was diagnosed with cardiac arrhythmia, and consequently implanted with pacemaker or cardioverter defibrillator. The patients treated with either oral anticoagulants or diuretics, but presumably also with ACE inhibitors or AT receptor blockers, had significantly higher odds for suffering from pacemaker-requiring arrhythmia. As to our best knowledge, this is the first study to show that the use of certain drugs affects diagnostic value of ILR in determining indications for pacemaker implantation.

The first use of ILR-like monitoring dates back to 1990, when Murdock and colleagues (19) performed a pilot study on 16 patients suffering from unexplained syncope, which were implanted with permanent pacemaker that had the capacity to record cardiac rhythm. Although markedly hindered by necessity for (and complications of) pacemaker implantation, even this first ILR prototype model clearly demonstrated usefulness in detecting or excluding cardiac arrhythmia as a cause of recurrent syncope, introducing itself as a new potential approach among other diagnostic methods. With technology development, the major limitations, such as requiring endocardial lead, being rather big in size, and not providing timely rhythm-event association, have been overcome (20), with modern ILR representing small, precise, and long-lasting device that is easy to insert and handle, and that poses low to no risk of any significant complications (21).

Over the last couple of decades, the utility of ILR in evaluation of unexplained syncope or palpitations has been often compared and almost invariably shown superior to other conventional methods (14). In one of the earliest randomized controlled studies, which included 60 patients with unexplained syncope and employed partial cross-over design, ILR-based 1-year-long monitoring strategy was deemed more likely to provide a diagnosis than short-term (up to 4 weeks) ECG monitoring followed by tilt and electrophysiological testing, with underlying arrythmias being detected by ILR and alternative strategy in 55% and 19% of the patients, respectively (3). Few years later, in 201 patients suffering from recurrent syncope, and randomized to either ILR or conventional investigation group, 6 months of follow-up revealed that the ECG diagnosis was made in 33% by former, as opposed to only 4% by latter approach (5). In a recent open-label, randomized controlled trial, involving 30 young patients with structurally normal heart but experiencing recurrent unexplained palpitations or syncope, 1 year of follow-up showed that the implantation of ILR provides significantly higher detection rate than conventional extended Holter monitoring (71.5% vs 18.7%) (14). High diagnostic yield of ILR has been confirmed by many other studies, ranging from 31%, documented by a multicenter, cohort study from Greece that included 871 patients implanted with ILR due to presyncope, syncope, cryptogenic stroke, or palpitations (8), to 65%, as reported by an observational, retrospective study analyzing data on 1360 German ILR recipients with a diagnosis of syncope (22). In our study, after 2 years of follow-up, cardiac arrhythmias were diagnosed by ILR in 36% of the patients, which conforms well to the previous reports, and indicates ILR real-world effectiveness. In line with many other similar studies (14, 18, 23), the most frequent cardiac arrhythmias we observed were pauses in cardiac activity and bradycardia.

According to the previous investigations, the early use of ILR in patients with unexplained syncope also results in lower healthcare-related costs compared with the conventional evaluation approach (5), especially if used in a population at higher risk of potentially dangerous, but treatable diseases (24). Based on the ESC classification, there are 5 major categories of syncope, of which cardiac arrhythmia-related type has been associated with the highest one-year overall mortality rate of up to 33% (16). Interestingly, of all the treatments prescribed to treat syncope, only cardiac pacing has been consistently associated with improvement of symptoms (5). As the management and the outcome of up to 90% of all ILR-detected symptomatic bradyarrhythmias largely depends on pacemaker implantation (25, 26), in the present study our main goal was to assess the diagnostic value of ILR in detecting arrhythmias requiring pacemaker implantation. As expected, the most frequent indications for a pacemaker insertion we detected were pauses in cardiac activity, followed by bradycardia. Our results comply with the findings of the most recent ILR-related systematic review and meta-analysis, which reported sinus node dysfunction (either sinus pause or sinus bradycardia) as the most common pacemaker-managed arrhythmias (18, 27).

Nevertheless, not all ILR recipients with bradyarrhythmia-related syncope seem to benefit equally from permanent pacemaker, so timely identification of pacemaker implantation predictors, combined with early ILR application, could reduce morbidity and mortality, as well as the cost of the treatment (28). So far, many factors have been reported to independently predict pacemaker requirement, including older age, obesity, diabetes, and hypertension (18, 27–30). However, in our study neither demographics (such as gender and age) nor medical conditions (comorbidities and ILR indications) were observed to significantly affect the odds of pacemaker implantation.

Regarding gender, our results are in line with most of the available studies (18), while rare observations of the opposite are usually considered as an incidence finding (28). However, in terms of age, the previous studies were almost completely consistent in classifying elderly patients (older than 65 or 75) as more likely pacemaker recipients (18, 31); as possible explanations they offered age-associated degenerative fibrosis of the cardiac conduction system, or other covariables usually associated with aging, such as polypharmacy, electrolyte imbalance, prior cardiac disease, or surgery. In our study, we did observe the tendency toward positive age-pacemaker correlation, yet we did not find it significant. We believe the discrepancy between our results and the majority of other reports most likely ensues from the difference in age structure of the patients (the medium age of our study group was <50 years, as opposed to others dealing with older populations (28-30, 32)), as well as from the small number of our study subjects belonging to the older age group. Although we are not completely unique in failing to correlate age with pacemaker requirement, It is worth noting that other studies with similar conclusions actually experience similar limitations (27). Therefore, we believe that adjusting the age structure of the population, together with increasing the population size, would likely improve the validity of the future studies and accuracy of their findings.

As for the comorbidities and ILR indications, the lack of association between any of them and permanent pacemaker implantation in our study population mostly corresponds to the available literature. Namely, in the recent systematic review and meta-analysis evaluating 8 different studies and 1007 ILR recipients (18), arterial hypertension has been associated with higher risk of pacemaker requirement. However, of 8 studies evaluated therein, 4 that covered European population differed in terms of predictive value of hypertension: 2 of them did not find it relevant (28, 30), while in other 2 its significance, detected by univariable analysis, was lost after controlling for other factors of influence (27, 32). Similarly, the history of diabetes does not seem to affect the odds of pacemaker requirement either (27, 28, 30), with only one study suggesting the opposite, and assuming low tolerance to bradycardia due to diabetic neuropathy to be the reason for their conclusion. Based on the available literature, the role of both arterial hypertension and diabetes in predicting the need for permanent pacemaker implantation thus seems unlikely, but, having in mind the overall prevalence of these diseases, further investigations are welcome. Other comorbidities and ILR indications investigated in our study were either not explored before (18), or did not prove to be important for prediction of pacemaker requirement in ILR recipients (27, 29, 30, 32).

Conversely, we have observed strong association between the use of certain drugs and the pacemaker insertion in patients diagnosed with cardiac arrhythmias. Namely, 3 out of 4 patients in our study were treated with at least one drug, with every tenth, every fourth, and almost every second being on the therapy with oral anticoagulants, diuretics, and ACE inhibitors or AT receptor blockers, respectively. Our results showed that the treatment od ILR recipients with any of these three drug groups is associated with higher odds of having pacemaker implanted. Our conclusions proved to be difficult to put into the context of other investigations, as there are not many previous studies dealing with the predictive value of concomitant medications in this regard.

In relation to renin-angiotensin system (RAS) inhibitors, their role as potential pacemaker implantation predictors was assessed, but no significant association was reported (30). It is well known that RAS inhibitors have little to no effect on heart rate in healthy subjects (33). However, when used in congestive heart failure (34), these drugs can improve vagal reactivity and lead to baroreflex-mediated bradycardia (35), supporting our observation of higher odds of pacemaker implantation in ILR recipients treated with ACE inhibitors or AT receptor blockers. Moreover, in specific circumstances such as bilateral renal artery stenosis, ACE inhibitors can cause a decline in renal function and renal failure (36, 37), which has been reported as an independent predictive factor for pacemaker implantation (27). Finally, these drugs are known to cause hyperkalemia (37, 38), which, if severe, can result in both bradycardia (39) and palpitations (40), contributing to symptomaticity of arrhythmias that demand pacemaker implantation. It should be mentioned that the association between the use of ACE inhibitors or AT receptor blockers and higher odds of pacemaker requirement, detected in our study, lost its significance after controlling for other factors of influence. Our observations are in line with the previous report by Huemer et al. (27), who identified the use of AT blockers as predictive of pacemaker implantation in ILR recipients, but their significance dropped after joint comparison with other more important factors of influence. Therefore, additional investigations are warranted to confirm the observed effect.

On the contrary, the influence of oral anticoagulants and diuretics, identified as predictive of pacemaker insertions in ILR recipients, remained significant even after multivariable analysis, becoming the only confirmed pacemaker predictors in our study. Regarding oral anticoagulants, both vitamin K- and non-vitamin K-antagonists were found to be regularly used by a number of study participants. There is no clear causal relation between oral anticoagulants and pauses in cardiac activity or bradycardia, as these drugs are not typically known to cause arrhythmias (41). However, one of the most common sustained cardiac arrhythmia, i.e. atrial fibrillation, represents an indication for the treatment with oral anticoagulants, due to their high efficacy in prevention of impending thromboembolism and the consequent embolic stroke (41). As atrial fibrillation often exhibit palpitations (42), and commonly occurs in patients with sinus node disfunction (43), we speculate that the higher pacemaker requirement among patients treated with oral anticoagulants could be attributed to coinciding symptomatic bradyarrhythmias.

In regard to diuretics, their use in relation to pacemaker implantation requirement was only sporadically investigated until now, with the results that mainly indicate the lack of any association. In a retrospective single center study by Huemer et al. (27) that involved 106 patients with ILR, diuretics were not identified as predictors of pacemaker implantation. The same conclusion was earlier drawn by Palmisano et al. (30), in a prospective multicenter study investigating 56 ILR recipients, with a follow-up of almost 2 years. However, it is well known that diuretics (except for potassium-sparing, which were not used by our study subjects) often lead to hypokalemia (44), due to excessive excretion of potassium as a “side effect” of their mechanisms of action (45). In the heart, potassium depletion causes reduction in repolarization reserve and intracellular Na^+^ and Ca^2+^ overload (45), which can generate wide variety of arrhythmias, including bradycardia (46). Therefore, we hypothesize that hypokalemia-related bradycardia could at least partly explain the predictive value of diuretic-based treatment for pacemaker implantation in ILR recipients. Although the studies on patients without ILR can not be fully comparable with those with ILR inserted, it is worth mentioning that the use of diuretics has been associated with pacemaker requirement in the presence of cardiac amyloidosis (47), while lower potassium levels were correlated with higher odds of permanent pacemaker implantation in patients with drug-related atrioventricular block (48), both suggesting the possibility of a similar effect in others suffering from conduction system impairment. Still, our study remains the first to detect the association between the use of oral anticoagulants and diuretics and permanent pacemaker implantation in ILR recipients. It should be noted, however, that it suffers from several limitations, including relatively small sample size, the lack of control group subjected to alternative diagnostic methods, and the lack of other demographic and medical data that might affect diagnostic efficacy of ILR and the pacemaker requirements.

In conclusion, our study shows that ILR represents an effective diagnostic approach in detecting cardiac arrhythmias requiring pacemaker implantation, especially in patients treated with oral anticoagulants or diuretics. The relevance of previous treatment with ACE inhibitors or AT receptor blockers remains to be confirmed in the future.

## Data Availability

Data will be available upon request.

## Acknowledgments

Authors are grateful to participants in the study.

